# Predicting Stroke and Mortality in Mitral Regurgitation: A Gradient Boosting Approach

**DOI:** 10.1101/2021.01.04.21249215

**Authors:** Jiandong Zhou, Sharen Lee, Yingzhi Liu, Tong Liu, Gary Tse, Qingpeng Zhang

**Author notes:** **Corresponding authors** Dr. Gary Tse, PhD, FESC, FACC, FHRS, FRCP, Tianjin Key Laboratory of Ionic-Molecular Function of Cardiovascular Disease, Department of Cardiology, Tianjin Institute of Cardiology, Second Hospital of Tianjin Medical University, Tianjin 300211, People’s Republic of China, P.R. China, Dr. Qingpeng Zhang, PhD, School of Data Science, City University of Hong Kong, Hong Kong, Hong Kong SAR People’s Republic of China.

## Abstract

**Introduction:** We hypothesized that an interpretable gradient boosting machine (GBM) model considering comorbidities, P-wave and echocardiographic measurements, can better predict mortality and cerebrovascular events in mitral regurgitation (MR).

**Methods:** Patients from a tertiary center were analyzed. The GBM model was used as an interpretable statistical approach to identify the leading indicators of high-risk patients with either outcome of CVAs and all-cause mortality.

**Results:** A total of 706 patients were included. GBM analysis showed that age, systolic blood pressure, diastolic blood pressure, plasma albumin levels, mean P-wave duration (PWD), MR regurgitant volume, left ventricular ejection fraction (LVEF), left atrial dimension at end-systole (LADs), velocity-time integral (VTI) and effective regurgitant orifice were significant predictors of TIA/stroke. Age, sodium, urea and albumin levels, platelet count, mean PWD, LVEF, LADs, left ventricular dimension at end systole (LVDs) and VTI were significant predictors of all-cause mortality. The GBM demonstrates the best predictive performance in terms of precision, sensitivity c-statistic and F1-score compared to logistic regression, decision tree, random forest, support vector machine, and artificial neural networks.

**Conclusion:** Gradient boosting model incorporating clinical data from different investigative modalities significantly improves risk prediction performance and identify key indicators for outcome prediction in MR.

## Introduction

Mitral regurgitation (MR), defined as a retrograde flow through the mitral valve during ventricular systole, is one of the most prevalent valvular heart disease worldwide with an estimated incidence of 1.7%[1-3]. The combination of aortic stenosis and MR accounts for three in four cases of valvular disease with an age-dependent increase in incidence of greater than 6% amongst those above the age of 65[3, 4]. Due to the chronic volume overload under MR, the left ventricular function deteriorates under remodeling, which ultimately leads to pulmonary hypertension and heart failure[5, 6]. The mechanisms underlying MR can organic, often degenerative, valvular defects, or functional problems secondary to left ventricular dysfunction [7]. Over the recent years, a variety of prognostic markers has been identified to improve the risk stratification in MR. Clinically, Besides from left ventricular ejection fraction, there is increasing evidence supporting the use of other electrocardiographic markers in outcome prediction, such as left ventricular end systolic diameter, peak mitral inflow velocity, and left atrial size [8-10]. Furthermore, P-wave indices, such as P-wave area and P-wave terminal force, were found to reflect left atrial remodeling and MR severity, hence may yield useful prognostic insights [11, 12]. In terms of laboratory markers, raised serum brain natriuretic peptide was found to associated with higher risk of cardiac event[13, 14]. The increase in inflammatory biomarkers, such as high-sensitivity C-reactive protein and raised neutrophil-to-lymphocyte ratio, were found to be associated with the adverse outcomes of MR, such as heart failure [15, 16].

Currently, there is yet a multi-parametric approach in the risk stratification of MR. Recently, we reported that risk stratification of MR can be significant improved with the use of a multi-task Gaussian process learning model which outperformed logistic regression [17]. In this study, we extend previous analyses by assessing the comparative performance of several machine learning models, such as Decision Tree (DT), Random Forest (RF), Support Vector Machine (SVM), Artificial Neural Network (ANN), and Gradient Boosting Machine (GBM).

## Methods

### Study population and baseline characteristics

This study was approved by the New Territory East Cluster-Prince of Wales Hospital (NTEC-PWH) Ethics Committee. The anonymized dataset on this study has already been made available in an online repository [18, 19]. This study include Han Chinese patients referred for echocardiography and subsequently diagnosed with MR during the period between 1^st^ March 2005 and 30^th^ October 2018. Comprehensive medical data were accessed from the healthcare database (Clinical Management System, CMS) that is linked to a territory-wide Clinical Data Analysis and Reporting System (CDARS) with unique reference identifier for each patient. Our team and other teams have previously used this system for epidemiological studies [20-22]. Clinical details including patient age, gender, blood pressure, smoking status, hypertension, diabetes mellitus, hypercholesterolemia, ischemic heart disease were extracted with the system. These characteristics and comorbidities were manually checked using CMS records to avoid under-coding. Automated hematological analyzer performed complete blood counts. Biochemical data including sodium, potassium, creatinine, urea, and albumin levels were also extracted. Neutrophil-to-lymphocyte ratio (NLR) was given by the ratio of peripheral neutrophil count/mm^3^ to peripheral lymphocyte count/mm^3^. The prognostic nutritional index (PNI) was calculated by 10 × serum albumin value (g/dl)□+□0.005 × peripheral lymphocyte count/mm^3^. Echocardiographic data was also obtained. Primary outcome is all-cause mortality, and secondary outcome is incident transient ischemic attack (TIA)/stroke.

### Electrocardiographic measurements

We extracted P-wave measurements of patients in sinus rhythm at baseline determined by electrocardiography and calculated the mean P-wave duration (PWD) from the leads V1, II, III and aVF. In addition, lead V1 was used to determine the amplitude of the P-wave for each included patient. Leads V1 to V6 as well as II, III and aVF were used to determine the P-wave morphology. P-wave duration (PWD) ≥ 120 ms in the absence and presence of biphasic P-waves in the inferior leads were used to indicate partial inter-atrial block (IAB) status and advanced IAB status of the patients. P-wave dispersion was determined according to the calculated maximum difference in PWD between the leads V1, II, III and aVF. P-wave terminal force in V1 (PTFV1) was determined as the area subtended by the terminal negative component of a biphasic P-wave in lead V1, and the area was calculated by the multiplication of the duration and waveform depth [23]. Abnormal PTFV1 was defined if it was > 40 ms.mV.

### Variable network analysis

One interesting exploration of the correlations between variables are the patterns of variable clustering, which then forms a variable network that can be visualized. In a variable network, each point represents a variable and each path represents a correlation between the two variables that it joins. The width and transparency of the path represent the strength of the correlation (wider and less transparent = stronger correlation). The positioning of variables can be handled by multidimensional scaling of the absolute values of the correlations. Variables that are more highly correlated appear closer together and are joined by stronger paths. Paths can also be colored by their sign (e.g., blue for positive and red for negative). The proximity of the points is determined using multidimensional clustering [24]. In this study, we first obtain the correlation matrix by calculating correlation coefficient of each variable pair, and then visualize the correlation matrix in a network diagram. In the diagram, each variable is represented by a node, and the connection between each pair of two nodes are shown by a colored path if the correlation reaches a threshold. The calculation and visualization are conducted by using the packages *igraph* and *corrr* in RStudio (Version 1.1.456).

### Interpretable gradient boosting learning approach

Gradient boosting machine (GBM) [25], a state-of-the-art machine learning method, was used to identify a set of key leading indicators that may help predict TIA/stroke and all-cause mortality. The idea behind boosting is that each sequential model builds a simple weak learner model to slightly improve the remaining errors. At each iteration, a new weak tree learner is trained with respect to the error the whole ensemble learnt so far. More details about GBM can be found in [25] and [26].

### Outcomes and statistical analysis

The primary outcome is all cause mortality, and secondary outcome is TIA/stroke. Evaluation metrics including precision, recall, and F1-score of using gradient boosting machine model were calculated and compared to benchmark models of logistic regression (LR), decision tree (DT), and the random forest model (RF). DT reveals from observational variables (represented in the branches) to target value (represented in the leaves), and was used for cardiovascular disease diagnosis, such as in-hospital mortality [27], congestive heart failure [28] etc. RF, first proposed by Breiman [29], is an ensemble approach for building predictive models where a forest is formed using a series of decision trees that act as “weak” learners. As individual trees, they are poor predictors, but can produce a robust prediction in aggregate form. Owing to its simple nature, lack of strong assumptions, and general high prediction performance, RF has been successfully used in many medical applications including prediction of severe asthma exacerbations [30], hospital readmissions in heart failure [31], non-invasive classification of pulmonary hypertension [32], etc. However, it should be noted that GBM is typically used with decision trees of a fixed size as base learners. RF combines results at the end of the process (by averaging or “majority rules”) while GBM combines results along the process. RF builds each tree independently while GBM builds one tree at a time. GBM as an additive model works in a forward stage-wise manner, introducing a weak learner to improve the shortcomings of existing weak learners. In addition, we also include black-box-like machine learning approaches of support vector machine (SVM) and artificial neural network (ANN) as baseline models for risk stratification. Statistical analysis was conducted using Stata (Version MP 13.0) and RStudio (Version 1.1.456). Experiments are simulated on a 15-inch MacBook Pro with 2.2 GHz Intel Core i7 Processor and 16 GB RAM.

## Results

### Baseline characteristics and network visualization of variables

A cohort of patients diagnosed with mitral regurgitation at a single tertiary centre (n=706; 57% male; median age: 66 [57-75] years old) was included in this study. Their clinical and laboratory parameters at baseline are shown in **Table 1**, stratified by cerebrovascular event outcome (*top*) or mortality outcome (*bottom*). The principles of the GBM model are illustrated in **Figure 1**, where we build an ensemble of shallow and weak successive tree-based learners, then sequentially combine a set of weak learners to deliver improved prediction accuracy. The network of variables constructed by the correlations between variables are shown in **Figure 2**, yielding patterns of variable clustering and the correlation strength of variable pairs. For instance, a cluster is formed by the highly correlated variable pairs of LVEF and LVESD, LVEDD and LVESD, LVDD and LVESD, and LVEDD and LVDD. Variables that are more highly correlated appear closer together and are joined by stronger paths. Blue and red colours denotes positive and red correlation, respectively. Strong correlations are observed between sex, urea, creatinine, regurgitant volume, and MR severity, LVDs.

**Table 1.** Baseline characteristics stratified by outcome of TIA/stroke (left) and all-cause mortality (right). Expressed as median (Q1-Q3) for continuous variables or frequency (percentage) for categorical variables. P-values indicate comparisons between the groups. Abbreviations are the same as those defined in the legend for Figure 1.

| Variable | No TIA/stroke | TIA/stroke | P-value | Alive | Dead | P-value |
| --- | --- | --- | --- | --- | --- | --- |
| Sex | 355<br>(355/613) | 48 (48/92) | 0.292 | 334<br>(334/587) | 69 (69/118) | 0.768 |
| Age | 66 (57-74) | 69 (57-77) | 0.286 | 64 (56-72.75) | 7 (66-81) | 1.1e-10 *** |
| LVH | 198<br>(198/613) | 31 (31/92) | 0.798 | 187<br>(187/587) | 42 (42/118) | 0.437 |
| SBP | 128 (113-144) | 130.5<br>(115.75-147.25) | 0.987 | 129 (115-144) | 125 (109.25-144.5) | 0.0633. |
| DBP | 73 (65-83) | 74 (64-81) | 0.088 | 74 (65-83) | 70 (61.25-83.75) | 0.389 |
| Smoking | 196<br>(196/613) | 28 (28/92) | 0.775 | 177<br>(177/587) | 47 (47/118) | 0.0383 * |
| Hypertension | 511<br>(511/613) | 79 (79/92) | 0.524 | 483<br>(483/587) | 107 (107/118) | 0.0224 * |
| Diabetes mellitus | 134<br>(134/613) | 24 (24/92) | 0.370 | 109<br>(109/587) | 49 (49/118) | 3.91e-08 *** |
| High Cholesterol | 164<br>(164/613) | 22 (22/92) | 0.553 | 141<br>(141/587) | 45 (45/118) | 0.00159 ** |
| IHD | 219<br>(219/613) | 32 (32/92) | 0.869 | 186<br>(186/587) | 65 (65/118) | 1.01e-06 *** |
| PSMR | 377<br>(377/613) | 48 (48/92) | 0.092 | 381<br>(381/587) | 45 (45/118) | 1.87e-08 *** |
| Severity | 609<br>(609/613) | 92 (92/92) | 0.759 | 583<br>(583/587) | 117 (117/118) | 0.674 |
| LVDd | 5.2 (4.7-5.8) | 5.2 (4.625-5.8) | 0.842 | 5.1 (4.6-5.8) | 5.4 (4.9-5.9) | 0.0335 * |
| LVDs | 3.5 (3-4.2) | 3.5 (3.1-4.35) | 0.850 | 3.5 (3-4.1) | 4.1 (3.2-4.9) | 0.2 |
| LADs | 4.2 (3.5-4.8) | 4 (3.4-4.6) | 0.200 | 4.1<br>(3.4925-4.7) | 4.6 (3.5-5.15) | 0.00356 ** |
| VTI | 156.8<br>(135.4-181.25) | 161.3<br>(145.4-188) | 0.952 | 157.4<br>(136.175-181.375) | 160.15<br>(136.625-190.55) | 0.815 |
| MR volume | 157.1<br>(63.675-332.9) | 148.2<br>(76.15-364.2) | 0.185 | 168.1<br>(66.225-342.75) | 122 (65-311.7) | 0.353 |
| ERO | 0.27 (0.16-0.435) | 0.25 (0.18-0.36) | 0.570 | 0.27 (0.165-0.44) | 0.23 (0.13-0.39) | 0.575 |
| MRVol | 41.995 (26.1-67.015) | 39.2 (27.75-59.25) | 0.372 | 42 (26.8-67.95) | 37 (24.4-54.1) | 0.0667. |
| LVEDD | 5.2 (4.7-5.8) | 5.2 (4.65-5.8) | 0.751 | 5.1 (4.6-5.8) | 5.4 (4.9-5.9) | 0.411 |
| LVESD | 3.5 (3-4.2) | 3.5 (3.1-4.4) | 0.514 | 3.49 (3-4.1) | 4.1 (3.225-4.975) | 5.34e-06 *** |
| LVEF | 59 (50-65) | 56.95 (41.75-64.05) | 0.0913 | 60 (52-65) | 50 (35-60) | 2.6e-08 *** |
| Symptomatic | 197 (197/613) | 24 (24/92) | 0.244 | 176 (176/587) | 45 (45/118) | 0.0817. |
| NYHA class | 610 (610/613) | 92 (92/92) | 0.231 | 575 (575/587) | 117 (117/118) | 0.0279 * |
| Surgery | 259 (259/613) | 35 (35/92) | 0.446 | 260 (260/587) | 34 (34/118) | 0.00183 ** |
| Average P-wave duration | 112.3 (97.625-124.225) | 107.5 (97-121.3) | 0.104 | 109.9 (96.8-123.5) | 113.5 (103.25-123.3) | 0.171 |
| Lymphocyte count | 1.3 (0.8-1.715) | 1.2 (0.9-1.6) | 0.753 | 1.4 (1-1.8) | 0.9 (0.7-1.5) | 5.69e-07 *** |
| Neutrophil count | 5.1 (3.8-7.1) | 5.61 (4-7.48) | 0.119 | 5.02 (3.8-7.2) | 5.5 (4-7.15) | 0.23 |
| Albumin | 36.9 (32-41.4) | 36.1 (30.7-40.3) | 0.316 | 37.9 (33.2-41.7) | 33.4 (28.65-37.8) | 9.29e-10 *** |
| Creatinine | 90 (72-117) | 94.5 (74.25-110.5) | 0.755 | 87 (71-106) | 117.5 (87-190.75) | 3.57e-15 *** |
| Platelet | 207 (166.5-263.5) | 202 (161.25-256) | 0.599 | 209 (171-269) | 197.5 (151.25-239.25) | 0.0427 * |
| Potassium | 4 (3.7-4.3) | 4.1 (3.775-4.325) | 0.721 | 4 (3.7-4.3) | 3.95 (3.575-4.3) | 0.124 |
| Sodium | 139.6 (137-141.4) | 139.3 (137.175-141.225) | 0.679 | 139.6 (137.3-141.5) | 138.75 (136.05-140.725) | 0.216 |
| Urea | 4.9 (6.5-8.47) | 6.47 (4.54-8.9075) | 0.966 | 6.15 (4.81-7.85) | 8.885 (5.175-17.2825) | <2e-16 *** |

**Figure 1.**
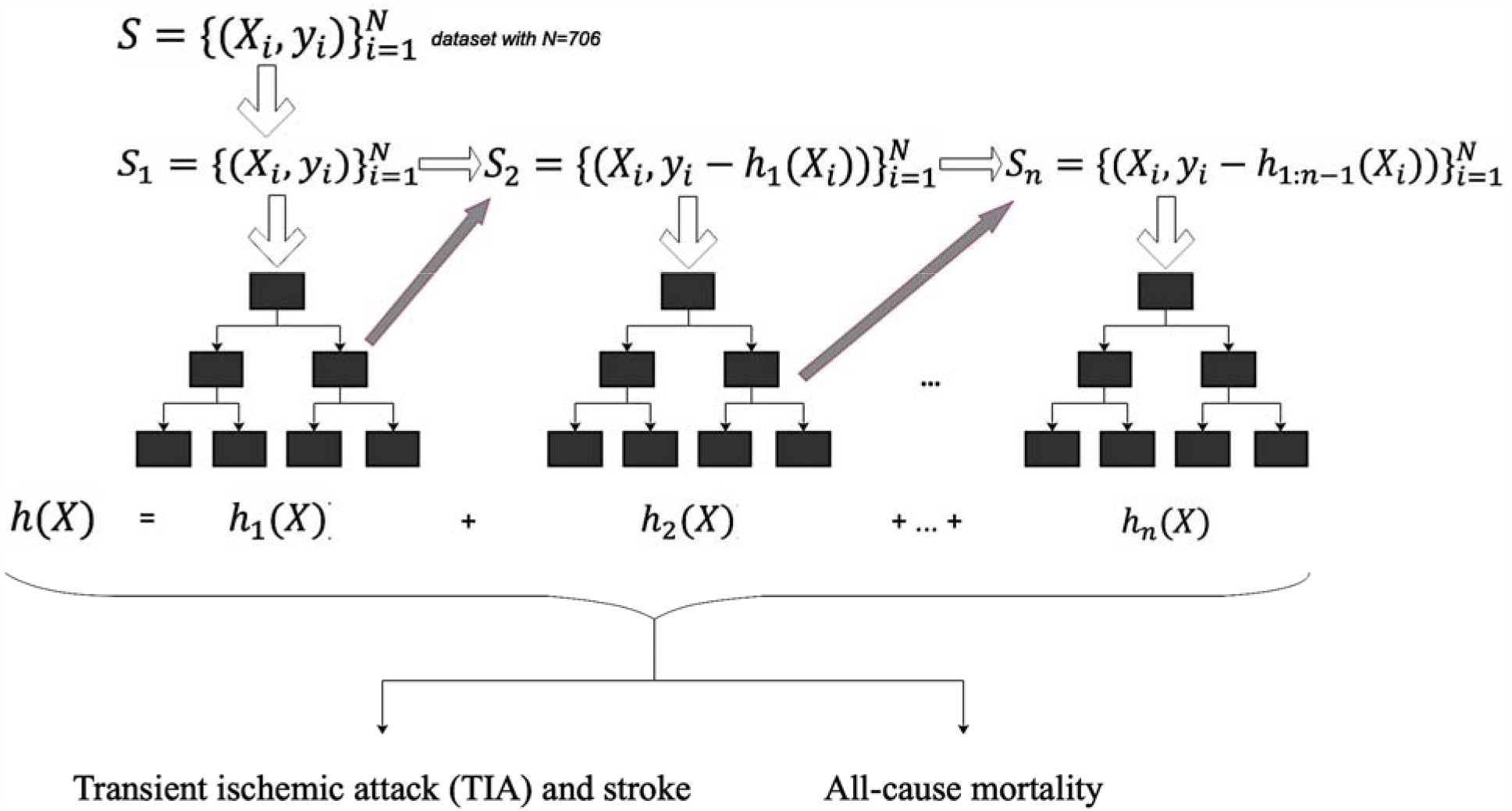
Diagram of sequential learning process of GBM prediction model.

**Figure 2.**
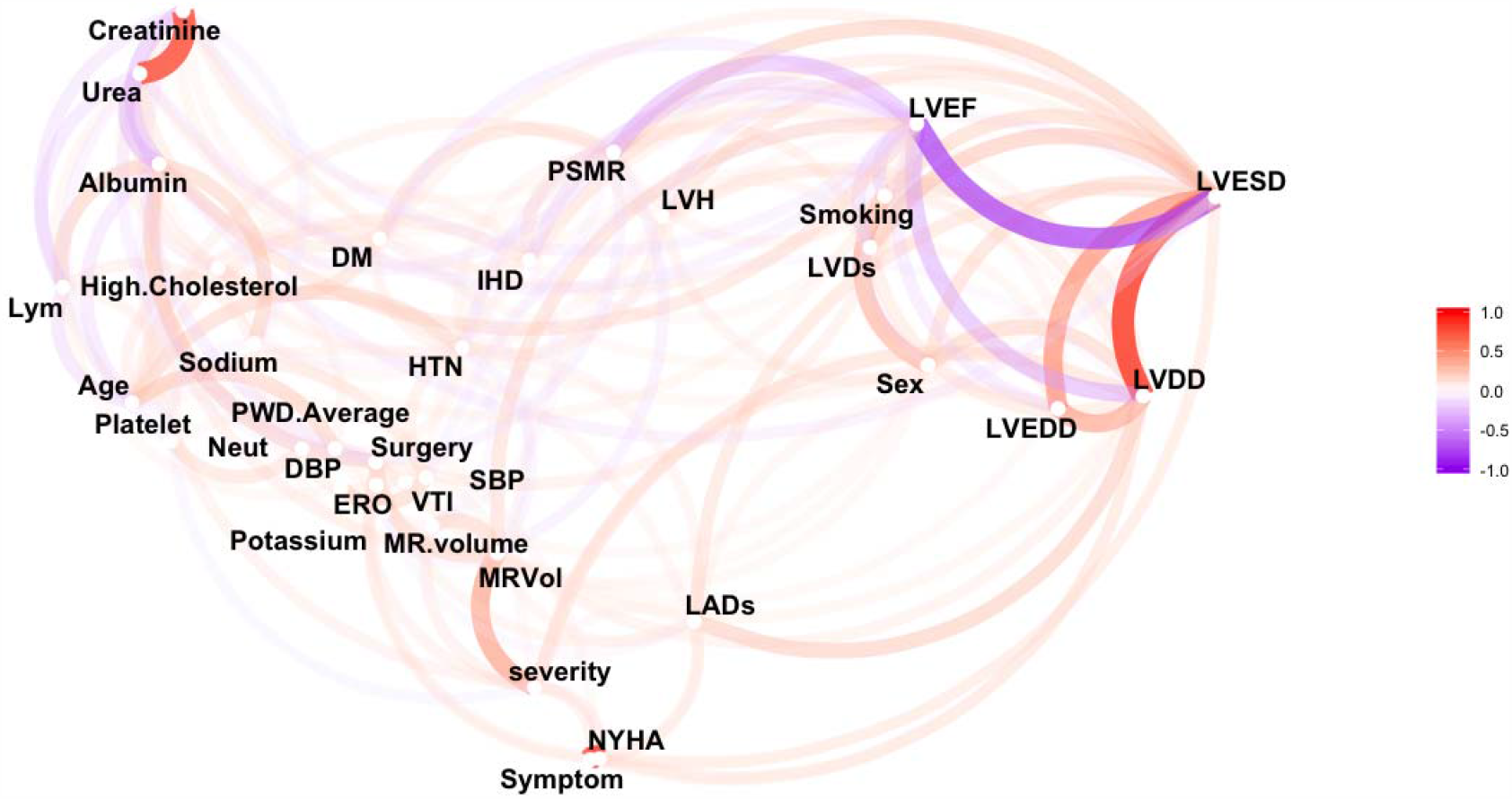
Variable correlation and clustering network. Abbreviations: LVH (left ventricular hypertrophy), SBP (systolic blood pressure), DBP (diastolic blood pressure), HTN (hypertension), DM (diabetes mellitus), PSMR (Primary or secondary mitral regurgitation), LVDs (left ventricular dimension at end systole), LVDd (left ventricular dimension at end diastole), LADs (left atrial dimension at end systole), VTI (velocity-time integral), ERO (effective regurgitant orifice), MRVol (mitral regurgitation volume), LVEDD (left ventricular end-diastolic diameter), LVESD (left ventricular end-systolic diameter), LVEF (left ventricular ejection fraction), NYHA class (New York Heart Association class for heart failure).

### Performance comparisons for risk stratification in mitral regurgitation

We compare the performance of using LR, DT, RF, SVM, ANN, GBM to predict i) TIA/stroke and ii) all-cause mortality. All the models were trained with 80% of patients and tested with five-fold cross-validation approach using the remaining 20% patients. The computation results were evaluated using the following metrics of recall, precision, F1-score and area under ROC curve (AUC) (**Table 2)**. With cross-validation approach, the GBM model significantly produced better risk prediction performance compared with other baseline models in predicting both cerebrovascular events and all-cause mortality. As the important hyperparameters in the GBM model, the number of trees and the tree depth were tuned to be 500 and 5, respectively. The hyper-parameter tuning process was essential to improve the predictive performance of GBM model. For the SVM model, the radial kernel parameters gamma and cost of constraints violation were tuned to 0.01, and 10, respectively. For the ANN model, the number of units in the hidden layer was set to 4, and the decay was set to 0.05. The observations about model performance are consistent with previous studies that GBM exhibited better predictions than SVM and mixture discriminant analysis in non-medical research domains [33, 34].

**Table 2.**
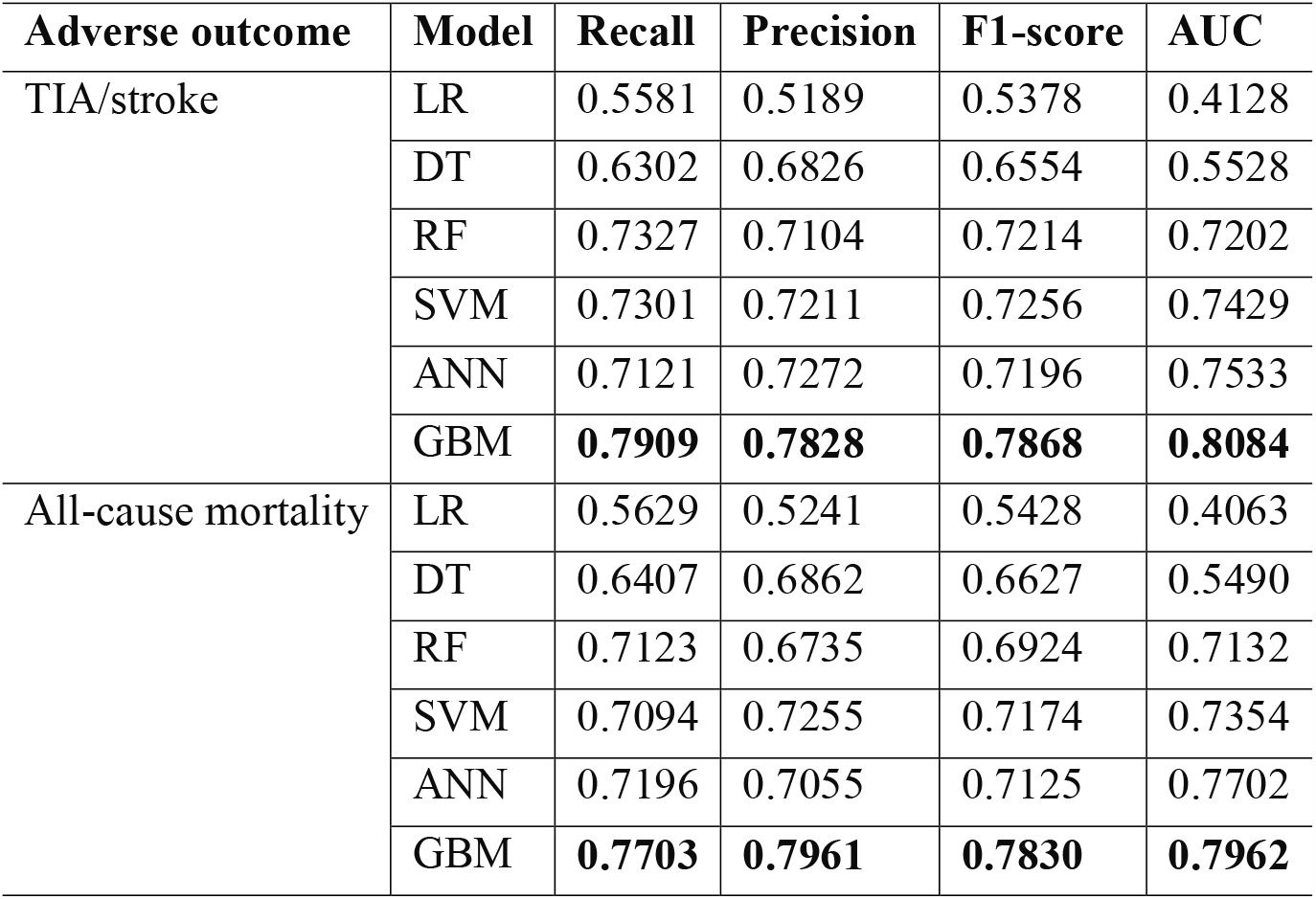
Performance comparison of Logistic Regression (LR), Decision Tree (DT), Random Forest (RF), Support Vector Machine (SVM), Artificial Neural Network (ANN), and Gradient Boosting Machine (GBM) in predicting TIA/stroke and all-cause mortality with five-fold cross-validation. The best metrics are shown in bold.

| Adverse outcome | Model | Recall | Precision | F1-score | AUC |
| --- | --- | --- | --- | --- | --- |
| TIA/stroke | LR | 0.5581 | 0.5189 | 0.5378 | 0.4128 |
|  | DT | 0.6302 | 0.6826 | 0.6554 | 0.5528 |
|  | RF | 0.7327 | 0.7104 | 0.7214 | 0.7202 |
|  | SVM | 0.7301 | 0.7211 | 0.7256 | 0.7429 |
|  | ANN | 0.7121 | 0.7272 | 0.7196 | 0.7533 |
|  | GBM | **0.7909** | **0.7828** | **0.7868** | **0.8084** |
| All-cause mortality | LR | 0.5629 | 0.5241 | 0.5428 | 0.4063 |
|  | DT | 0.6407 | 0.6862 | 0.6627 | 0.5490 |
|  | RF | 0.7123 | 0.6735 | 0.6924 | 0.7132 |
|  | SVM | 0.7094 | 0.7255 | 0.7174 | 0.7354 |
|  | ANN | 0.7196 | 0.7055 | 0.7125 | 0.7702 |
|  | GBM | **0.7703** | **0.7961** | **0.7830** | **0.7962** |

### Key predictors of adverse outcomes with GBM model

The GBM model calculates the importance (predictive strength) of variables to predict TIA/stroke (**Figure 3**, top) and all-cause mortality (**Figure 3**, bottom) in MR. The top ten most important variables for risk stratification of TIA/stroke and all-cause mortality are listed in **Table 3**. Average PWD, albumin, MR regurgitant volume, left ventricular ejection fraction (LVEF), left atrial dimension at end systole (LADs), velocity-time integral (VTI) and effective regurgitant orifice (ERO) play critical roles in predicting TIA/stroke, in descending order of importance. For all-cause mortality, urea, LVEF, platelet count, LADs, VTI, albumin, age, sodium, average PWD and LVDs are the most powerful predictors. Clearly, the optimum set of variables for predicting each outcome is different. For instance, average PWD is the most important predictor in the TIA/stroke model, while it is less important in the all-cause mortality model. By contrast, age was the most important predictive factor for all-cause mortality. In addition, we can find that variables that are highly correlated with those that show high predictive power may not show similar strong predictive strength.

**Table 3.**
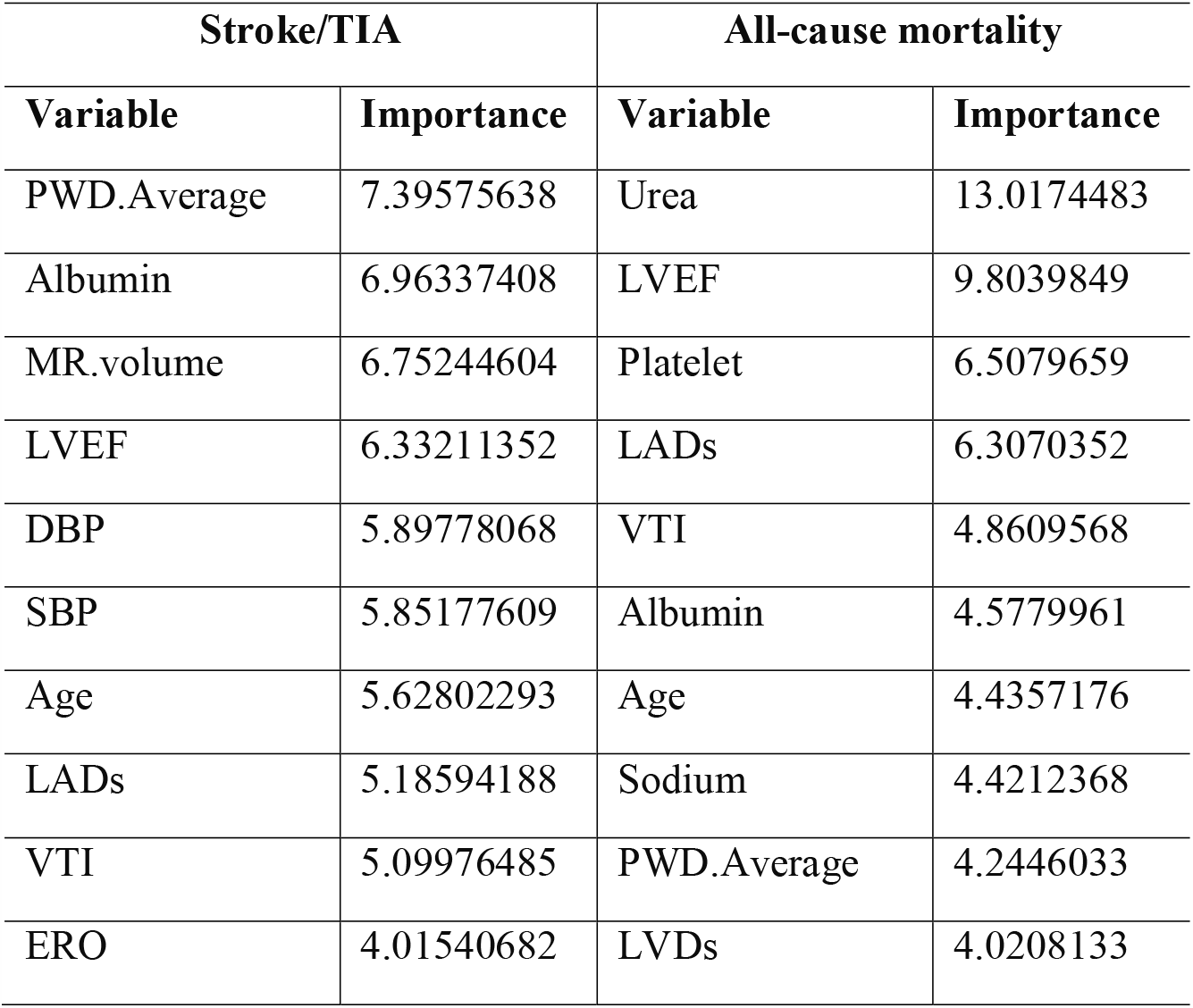
Top ten most important variables for GBM to predict stroke/TIA and all-cause mortality.

**Figure 3.**
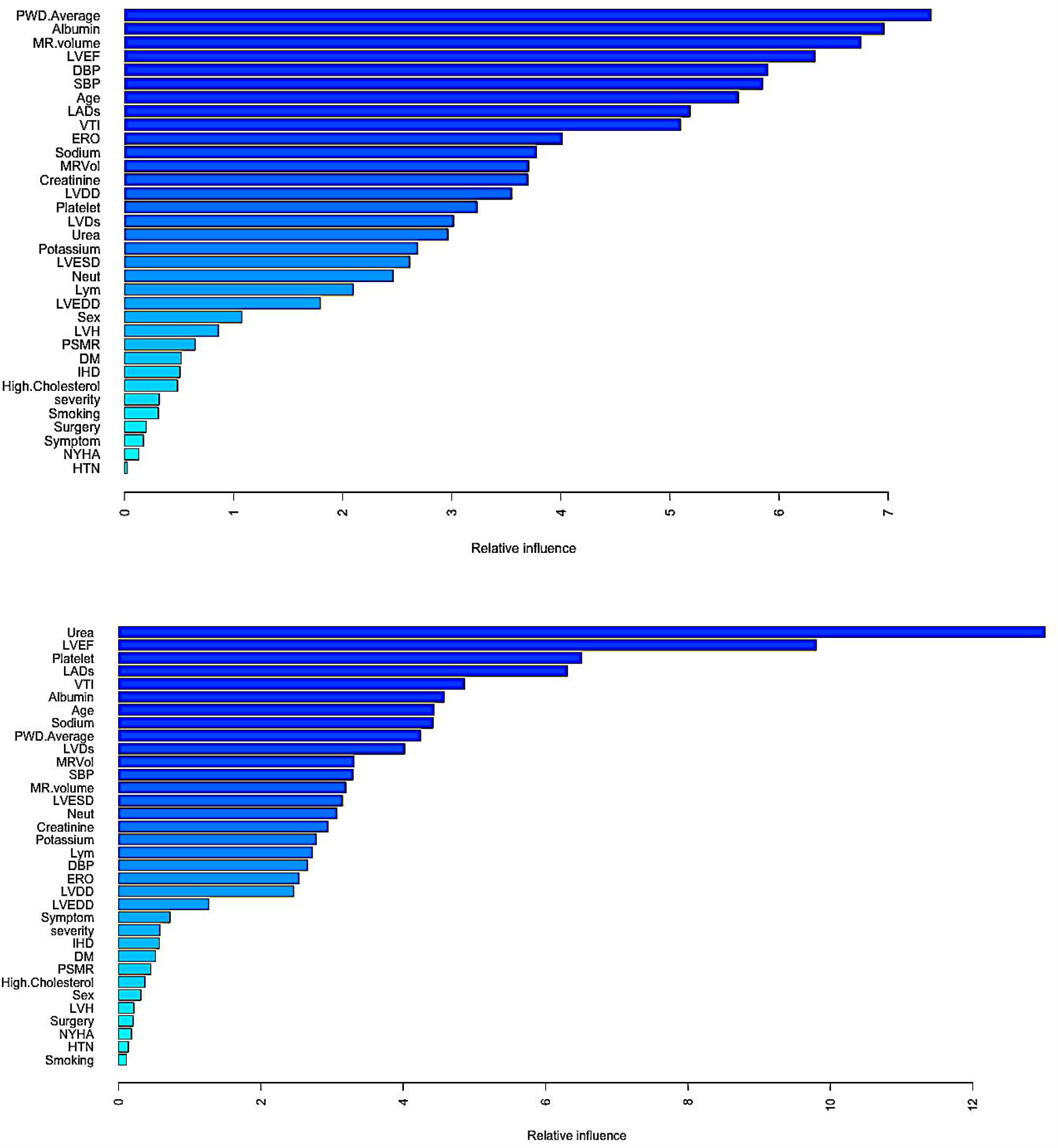
Variable importance plot for GBM to predict TIA/stroke (top) and all-cause mortality (bottom).

### Partial dependence of key risk stratification variables

Partial dependence plots generated by GBM provide additional insights on how the variables affect the adverse outcome. The partial dependence plots of the top eight most important variables for the stroke/TIA prediction model is shown in **Figure 4**. The deciles of the distribution of the corresponding variable are shown by the log odds and the hash marks at the base of each plot. The partial dependence of each predictor accounts for the average joint effect of the other predictors in the model. Average PWD, LVEF, albumin, and age have a nonmonotonic partial dependence. They decrease over the middle range and increases nearly at the highest values. MR Volume increases sharply before reaching 150 ml, decreases before 350 ml and then increases again at the end. For SBP, the risk fluctuates before reaching 170 mmHg and then abruptly increases at the end, followed by a small decrease. DBP has a roughly monotonically decreasing partial dependence followed by a long plateau still the end. Note that these plots are not necessarily smooth, since no smoothness constraint was imposed on the fitting.

**Figure 4.**
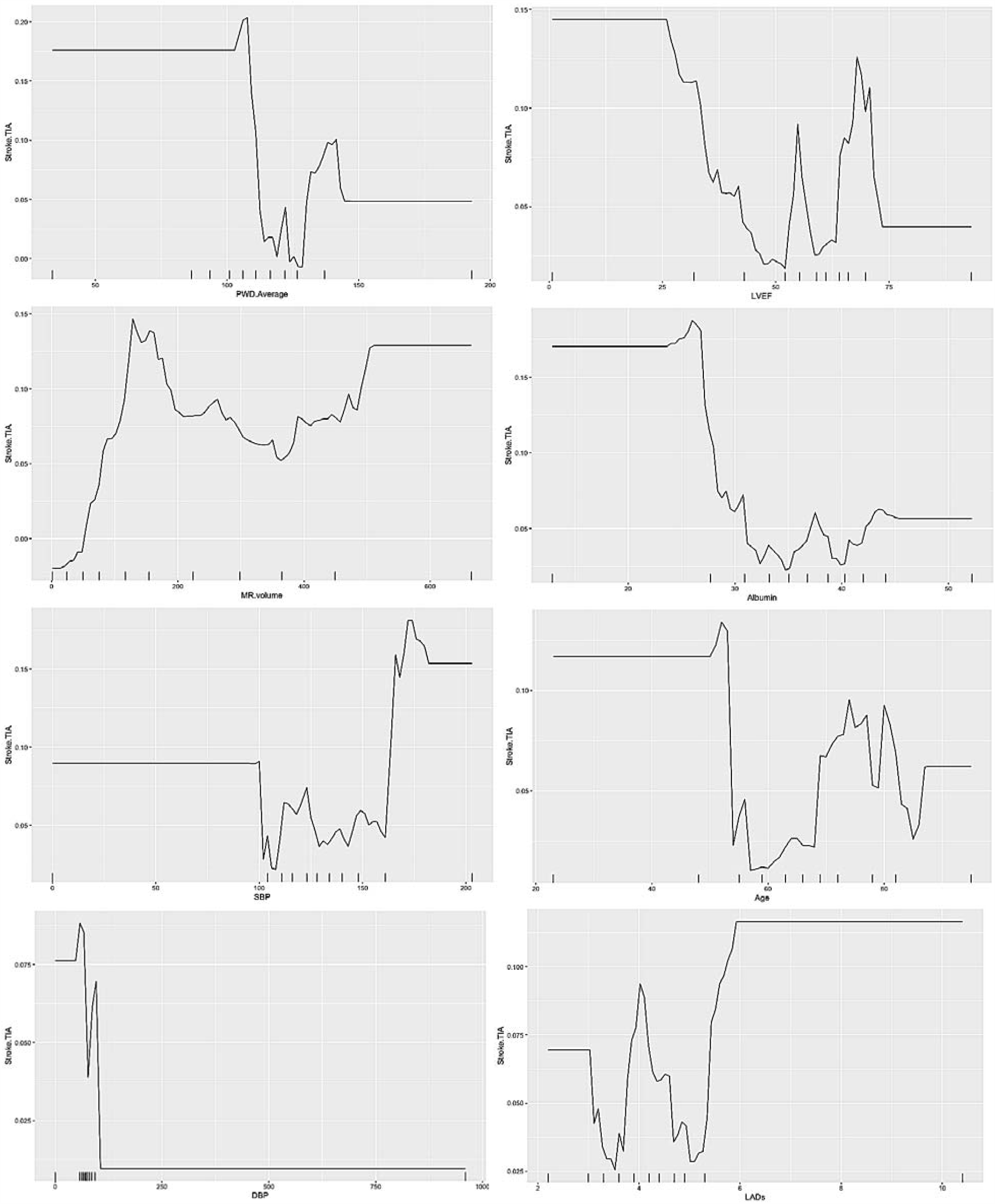
Partial dependence of six most important variables in predicting Stroke/TIA

The partial dependence plots generated by GBM between the different variables and all-cause mortality are shown in **Figure 5**. For both urea and LADs, there is a monotonic increase in the risk of mortality as their levels increase. The relationship between LVEF and mortality is complex, with mortality increase as LVEF decreases below 52%. Platelet and albumin show similar roughly monotonically decreasing partial dependence except an increase in the middle range levels. At very low VTI values, VTI decreases sharply with increasing mortality. Finally, for average PWD, there appears to be a U-shaped relationship with al-cause mortality.

**Figure 5.**
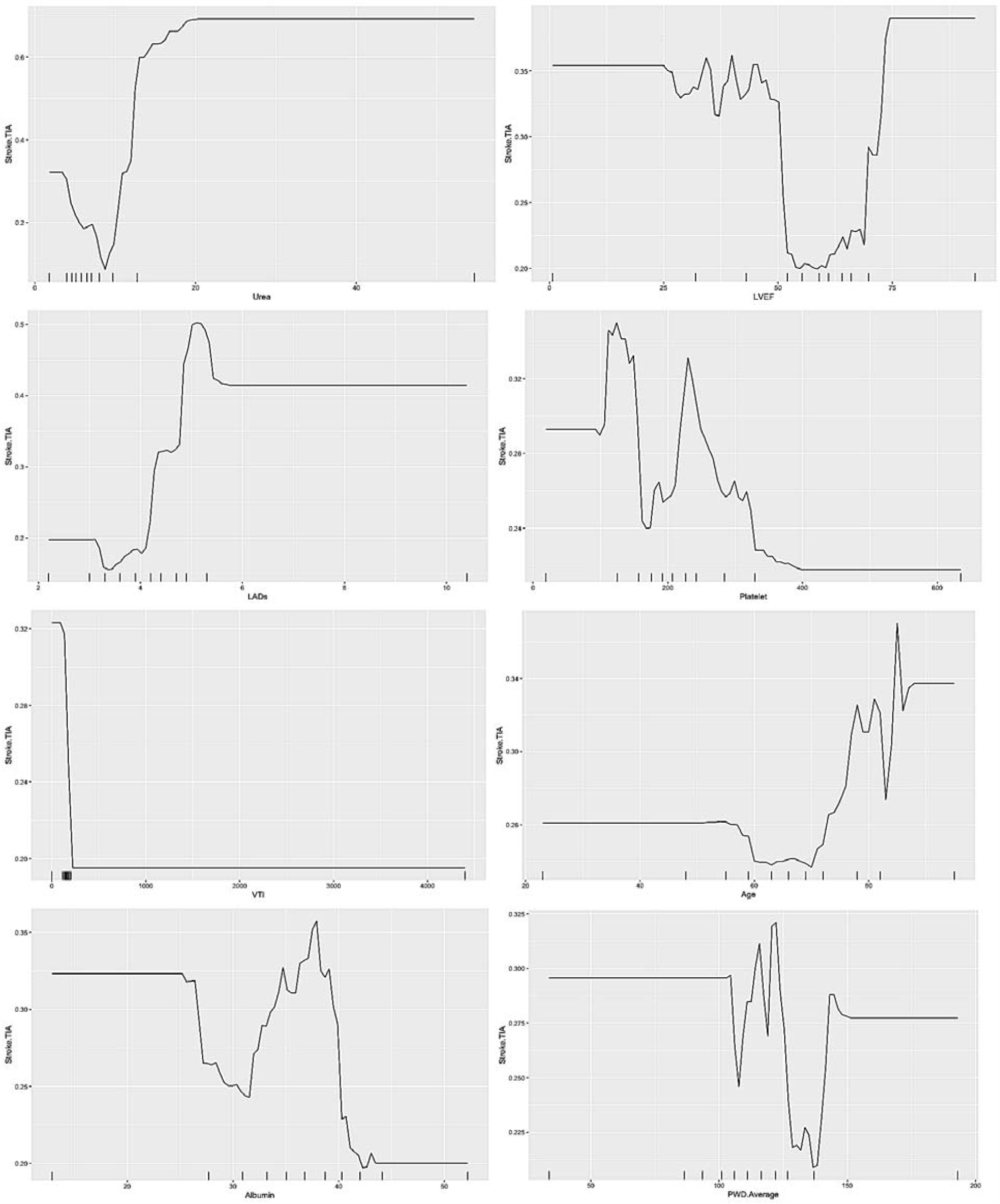
Partial dependence of eight most important variables in predicting all-cause mortality.

## Discussion

In this study, we found that an interpretable machine learning method with the consideration of baseline comorbidities, laboratory examinations reflecting inflammatory and nutritional states, electrocardiographic P-wave as well as echocardiographic measurements can accurately predict cerebrovascular and mortality in MR. Gradient boosting machine (GBM) significantly outperformed other approaches of logistic regression (LR), decision tree (DT), random forest (RF), support vector machine (SVM) and artificial neural network (ANN).

### A gradient boosting machine (GBM) method outperformed other machine learning techniques

GBM [25], a state-of-the-art machine learning method, was used to identify a set of key leading indicators that may help predict TIA/stroke and all-cause mortality. Previously, GBM models have been successfully used for MiRNA-disease association prediction [35], blood pressure prediction [36], and identification of medication relations with adverse drug events [37]. GBM generally showed better predictive performance in a series of model comparisons compared to other machine learning algorithms such as SVM and ANN [38]. Another study used a territory-wide database to predict stroke outcome and investigated the performance of SVM, ANN and also random survival forest [39], but did not compare it to a GBM approach. Another study using the United Kingdom General Practice database compared RF, LR, GBM and neural networks for first cardiovascular event in patients initially free from cardiovascular diseases [40]. GBM slowly but steadily achieves optimization by growing a series of weak decision trees in a stage-wise fashion which efficiently utilize the strengths of classification/regression trees and boosting. The superior predictive performance of the GBM model against the conventional models highlights the power of machine learning techniques in accounting for the intervariable nonlinear correlations, whilst maintaining interpretability, in outcome prediction. GBM is appropriate for risk stratification in the present study since it can improve the overall predictions by capturing the non-linearity in sparsely populated data. Besides from the advantages of quick convergence and accuracy improvement, GBM avoid overfitting since it can stop learning as soon as overfitting has been detected, typically by using cross-validation.

MR, a classical example of cardiovascular diseases where several factors interplay in its progression, is an ideal model for the application of machine learning. Although no other studies were noted to use machine learning in the risk stratification of MR, it has been applied in other cardiovascular diseases. For example, DT has been successfully used for both diagnosis [28] and prognosis prediction [27, 41]. Random forests, first proposed by Breiman [29], is an ensemble approach for building predictive models. RF has been successfully used in many medical applications including prediction of severe asthma exacerbations [30], hospital readmissions in heart failure [31], non-invasive classification of pulmonary hypertension [32]. Recently, RF was shown to demonstrate stronger predictive power in identifying predictors for the heterogeneity in response against pharmacotherapy amongst a large cohort of heart failure patients [42]. GBM is typically used with decision trees of a fixed size as base learners. RF combines results at the end of the process, by averaging or using “majority rules”, whereas GBM combines results along the process. RF builds each tree independently while GBM builds one tree at a time. GBM as an additive model works in a forward stage-wise manner, introducing a weak learner to improve the shortcomings of existing weak learners.

With increasing evidence supporting the superiority of machine learning in predictive accuracy, it is increasingly applied in multi-parametric risk stratification models. Jamthikar *et al*. developed a novel model based on RF that incorporates conventional risk factors with predictive features from carotid ultrasound image as an inexpensive and effective tool for cardiovascular/ stroke risk prediction[43]. Similarly, the WATCH-DM risk score was developed to predict the risk of incident heart failure during hospitalization amongst diabetic patients. The RF and DT based multiparametric score, which included clinical, laboratory and electrocardiographic variables, demonstrated better discrimination than the best-performing Cox-based model, which illustrate the potential in the incorporation of machine learning into clinical practice[44].

The prognostic use of markers found by the present study is well supported. Clinical predictors, such as age, diastolic and systolic blood pressure, were well established, and justified by their relation to the pathogenesis and disease progression of MR[45, 46]. For laboratory markers, uric acid has been found to correlate with left ventricular remodeling, MR severity and the outcome of heart failure in MR[47]. Serum albumin was reported to be lower in those with persistent MR after acute rheumatic fever[48]. Mean platelet volume, which reflects the platelet production and function, is associated with MR severity and thromboembolism risk[49, 50]. In terms of electrocardiographic markers, reduced LVEF, increased left atrial dimensions and left ventricular end diastolic diameter have been found to predict MR severity, which is associated with increased in-hospital cardiac death risk and overall mortality[47, 51, 52]. P wave indices reflective of left atrial remodeling, such as P wave area and P wave terminal force, was predictive of MR severity[11, 12]. In this study, P-wave duration was shown to be one of the most important predictors of incident stroke and was incorporated for risk prediction in our machine learning models.

### Study limitations

Several limitations should be noted. Firstly, it is limited by its retrospective nature and single ethnicity of the patients included. Secondly, data on some widely used clinical prognostic markers, such as results on exercise tolerance test, were not available for all patients. Finally, only the impact of medical or surgical treatment was not assessed in this study.

## Conclusion

An interpretable machine learning risk stratification model considering multi-modality clinical data can better predict cerebrovascular events and mortality in MR. Experiments demonstrate the advantage of GBM to significantly improve the overall risk stratification performance over baseline models, including LR, DT, RF, SVM, and ANN, in addition to provide good model interpretability about the predictive strengths of predictors. Partial dependences are also observed, which benefit insightful understanding on the effects of these predictive variables upon the adverse outcomes.

## Data Availability

Data available upon request

