## Supplementary Appendix for "Predicting Stroke and Mortality in Mitral Regurgitation: A Gradient Boosting Approach"

**Supplementary Table 1.** All variables based on their importance for classifying TIA/stroke (left) or all-cause mortality (right).

| **Stroke/TIA** | | **All-cause mortality** | |
| --- | --- | --- | --- |
| **Variable** | **Importance** | **Variable** | **Importance** |
| PWD.Average | 7.39575638 | Urea | 13.0174483 |
| Albumin | 6.96337408 | LVEF | 9.8039849 |
| MR.volume | 6.75244604 | Platelet | 6.5079659 |
| LVEF | 6.33211352 | LADs | 6.3070352 |
| DBP | 5.89778068 | VTI | 4.8609568 |
| SBP | 5.85177609 | Albumin | 4.5779961 |
| Age | 5.62802293 | Age | 4.4357176 |
| LADs | 5.18594188 | Sodium | 4.4212368 |
| VTI | 5.09976485 | PWD.Average | 4.2446033 |
| ERO | 4.01540682 | LVDs | 4.0208133 |
| Sodium | 3.77698651 | MRVol | 3.3096132 |
| MRVol | 3.70517901 | SBP | 3.2958917 |
| Creatinine | 3.69902418 | MR.volume | 3.1938672 |
| LVDD | 3.55146122 | LVESD | 3.1480428 |
| Platelet | 3.23426211 | Neut | 3.0699112 |
| LVDs | 3.017986 | Creatinine | 2.9433226 |
| Urea | 2.96505662 | Potassium | 2.7820843 |
| Potassium | 2.68866058 | Lym | 2.7265421 |
| LVESD | 2.6143291 | DBP | 2.6565923 |
| Neut | 2.46326905 | ERO | 2.5363241 |
| Lym | 2.09879985 | LVDD | 2.4635348 |
| LVEDD | 1.79634823 | LVEDD | 1.2690625 |
| Sex | 1.07719745 | Symptom | 0.7236196 |
| LVH | 0.86224307 | severity | 0.5835935 |
| PSMR | 0.64886875 | IHD | 0.576152 |
| DM | 0.5210202 | DM | 0.5208016 |
| IHD | 0.50870456 | PSMR | 0.4555814 |
| High.Cholesterol | 0.48659025 | High.Cholesterol | 0.3732136 |
| severity | 0.31966262 | Sex | 0.3145315 |
| Smoking | 0.310781 | LVH | 0.2165427 |
| Surgery | 0.19868107 | Surgery | 0.207654 |
| Symptom | 0.1746954 | NYHA | 0.1846737 |
| NYHA | 0.13188618 | HTN | 0.1401485 |
| HTN | 0.02592372 | Smoking | 0.1109409 |
